# Towards Democratizing CNS Tumor Classification: 10-fold reduction in methylation sequencing cost with the Flongle Flow Cell

**DOI:** 10.64898/2026.09.03.26361431

**Authors:** Assaf Grunwald, Galina Feinberg-Gorenshtein, Helen Toledano, Yehudit Birger, Shai Izraeli, Yuval Ebenstein

**Affiliations:** Department of Physical Chemistry, School of Chemistry, Tel Aviv University, Tel Aviv, Israel; Felsenstein Medical Research Center, Tel Aviv University, Petach Tikva, Israel; The Rina Zaizov Division of Pediatric Hematology-Oncology, Schneider Children’s Medical Center of Israel, Petach Tikva, Israel; Neuro-Oncology Unit, Dept. of Pediatric Hematology-Oncology, Schneider Children’s Medical Center, Petah Tikva, Israel; The Gray Faculty of Medical and Health Sciences, Tel Aviv University, Tel Aviv, Israel; Faculty of Medicine, Tel Aviv University, Tel Aviv, Israel; Department of Biomedical Engineering, Faculty of Engineering, Tel Aviv University, Tel Aviv, Israel

**Keywords:** CNS tumor classification, DNA methylation profiling, Oxford Nanopore Technologies

## Abstract

Oxford Nanopore Technologies-based methylation profiling enables rapid, accurate CNS tumor classification but is currently mostly performed with MinION flow cells (~$1,000 USD). We benchmarked the low-cost Flongle flow cell (~$100, 10-fold reduction) for methylation-based tumor classification. Across 12 pediatric CNS tumor samples with highly variable sequencing yields (17–330 Mbp), both Sturgeon and nanoDx classifiers achieved perfect diagnostic accuracy when operating above significance thresholds, despite extreme data sparsity. Bootstrap analysis determined empirically defined minimum data thresholds. These results demonstrate that Flongle flow cells enable cost-effective deployment of rapid, accurate nanopore-based CNS diagnostics while maintaining clinical reliability.

---

DNA methylation profiling using Oxford Nanopore Technologies (ONT) is increasingly being incorporated into clinical neuro-oncology workflows for rapid classification of central nervous system (CNS) tumors^1^. This approach builds on the reference methylation cohort established by Capper et al.^2^, which has been integrated into the World Health Organization (WHO) Classification of CNS Tumors as a key framework for resolving tumor types^3^. ONT-based classification has shown utility across a wide range of specimen types, including fresh frozen, intraoperative biopsies and degraded formalin-fixed, paraffin-embedded (FFPE) sample^4–8^.

Current clinical implementations generally use ONT MinION flow cells, which typically generate sufficient data for diagnosis within approximately two hours of sequencing and sequencing coverage of less than 1X; hence, the full potential data capacity of the MinION flow cell is not fully utilized. This is enabled by classification tools such as Sturgeon and nanoDx, which are designed to perform reliably under sparse CpG coverage^9,10^. More recently, M-PACT, a deep neural network-based classifier, was developed to impute missing CpG values specifically within cell-free DNA (cfDNA), offering a potential computational framework to handle even more extreme scenarios of data sparsity^11^.

While the ONT MinION sequencer is a highly deployable platform, its standard flow cells remain relatively expensive (~$1,000 USD), limiting scalability and broader clinical adoption. ONT also offers a smaller-format Flongle flow cell, a substantially lower-cost alternative (~$100 USD) by scaling down the active pore count (around 126 nominal pores compared to around 2048 in the MinION flow cell). Because the underlying sequencing chemistry and algorithmic frameworks remain fundamentally identical across both formats, the key operational shift on the Flongle is simply a lower rate and volume of data accumulation.

Whether existing classifiers remain reliable under these conditions is not fully established, having only been briefly noted in prior work without detailed evaluation^12^. Here, we benchmark Flongle-based CNS tumor classification across cumulative sequencing depth to assess how classification confidence evolves in time, and whether the Flongle presents a viable low-cost alternative for CNS tumor classification.

This benchmarking study evaluated a cohort of 12 pediatric CNS tumor samples spanning distinct histopathological entities **(Supplementary Table 1)**, conducted in accordance with the Declaration of Helsinki and approved by our local institutional Helsinki Committee (Supporting Information). Genomic DNA was isolated from fresh-frozen or FFPE tumor tissue collected during standard surgical intervention, using previously published protocols (Supplementary Information). Library preparation for methylation-aware sequencing was performed using the ONT Rapid Sequencing Kit (SQK-RAD114), bypassing multi-step ligation cleanups to maximize the conservation of limited low-input pediatric tumor derived DNA. Minor adjustments were introduced to the standard library preparation protocol to optimize sequencing yields from low-input DNA (Supporting Information).

Initial active pore counts ranged from 49–82 per flow cell **(Supplementary Table 1)**. Over a 24-hour run or until pore exhaustion, cumulative yield ranged from 17 to 330 Mbp *(Figure 1, Supplementary Table 1)*. This highly variable throughput provided an ideal empirical dataset to stress-test the tolerance of downstream methylation classifiers under conditions of extreme data sparsity.

**Figure 1.**
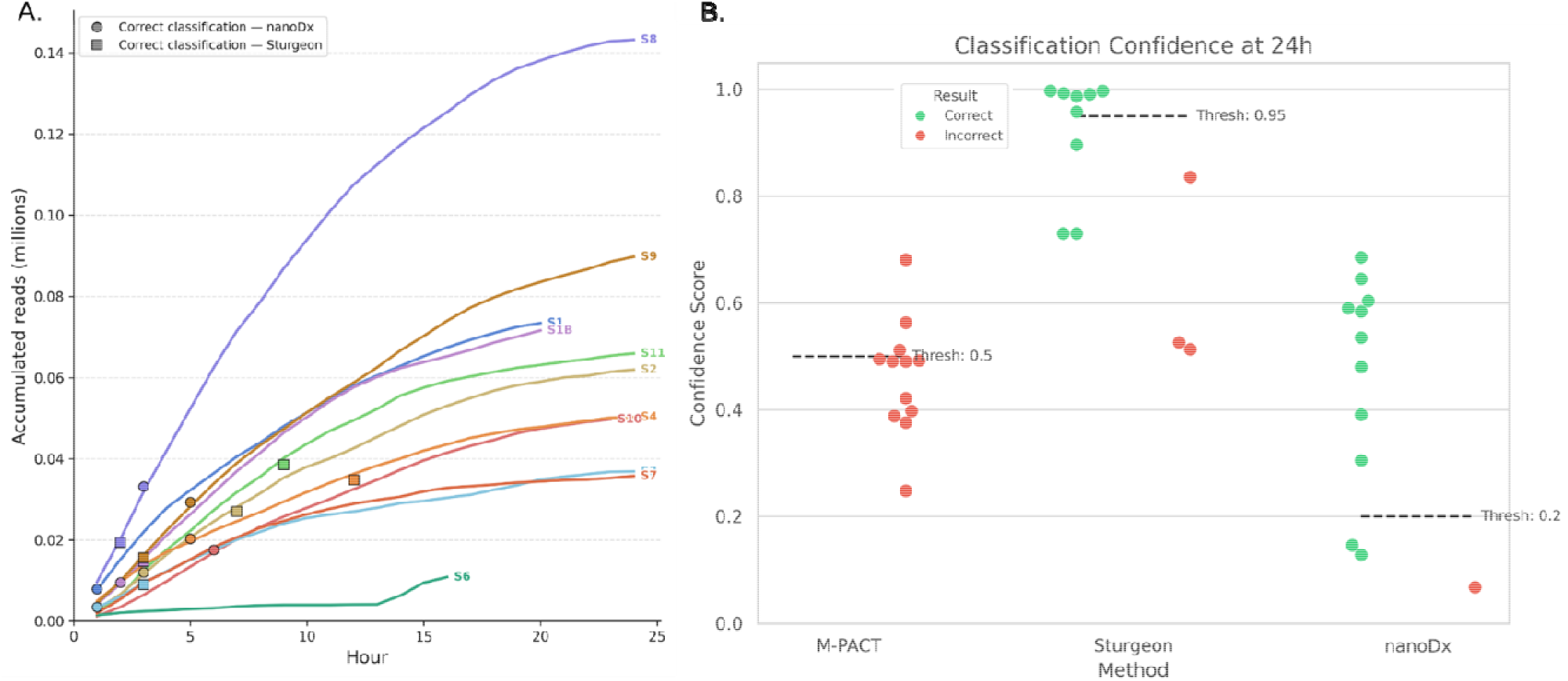
CNS tumor classification using Flongle nanopore sequencing. (A) Accumulated sequencing reads over time for each sample (colored lines). Circles (⍰) and squares (▪) indicate the first timepoint at which nanoDx and Sturgeon, respectively, reached a correct classification above their significance threshold. Samples that did not reach significant classification are not marked. (B) Classification confidence scores at 24 hours of sequencing for M-PACT, Sturgeon, and nanoDx. Each dot represents one sample, colored by classification outcome (green: correct, red: incorrect). Dashed lines indicate the classifier-specific confidence thresholds (M-PACT: 0.5, Sturgeon: 0.95, nanoDx: 0.2).

Subjecting this 12-run cohort to real-time algorithmic evaluation revealed marked architectural differences in how these models handle sparse data (**Figure 1B**). Across all 12 runs, the cell-free DNA-optimized M-PACT framework demonstrated severe instability, yielding outputs that were entirely discordant relative to the neuropathologist-confirmed ground truth. We therefore excluded M-PACT from downstream cohort evaluations to focus exclusively on the head-to-head performance of Sturgeon and nanoDx.

In this head-to-head comparison, both pipelines achieved pristine diagnostic accuracy at their final endpoints, with zero false classifications passing their respective significance thresholds **(Figure 1B)**. Specifically, nanoDx successfully delivered a statistically significant, correct classification in 75% of the cohort (9/12 runs), while the remaining cases failed to clear its significance barrier. Conversely, while the Sturgeon classifier does not define a fixed internal significance cutoff, applying its recommended stringent threshold (0.95) to ensure maximum diagnostic safety yielded correct, high-confidence classifications for the 6 samples that managed to clear this barrier (6/12 runs) **(Figure 1B, SI Table 1)**. While this conservative approach eliminates false positives, it carries the operational trade-off of leaving lower-yield, data-sparse samples unclassified. Given the extreme data constraints and pore volatility inherent to Flongle flow cells, it is highly remarkable that every single statistically significant classification across both architectures aligned perfectly with the neuropathologist-confirmed ground truth.

To define the minimum data requirements for confident diagnostics, we first evaluated the relationship between sequencing time, cumulative throughput, and the milestone at which each classifier achieved a correct and statistically significant assignment **(Figure 1A)**. Unsurprisingly, because starting DNA inputs and initial active pore counts inherently vary between individual low-cost flow cells, real-time data accumulation rates exhibited high sample-to-sample volatility **(Figure 1A)**. An evaluation of classifier confidence as a direct function of absolute data volume is shown in supplementary Figure 1.

Classification as a function of accumulated data is illustrated by two contrasting, representative cases **(Figure 2 A, B)**. For the more challenging Sample S1 (Embryonal - CNS NB - FOXR2), early data scarcity led Sturgeon to produce transient, discordant assignments (red dots); however, as data accumulated, these incorrect outputs resolved into stable, correct, yet sub-threshold assignments (green dots). In contrast, nanoDx crossed and maintained its significance threshold from the earliest checkpoints **(Figure 2A)**. Conversely, for the best performing sequencing run of Sample S8, both classifiers achieved rapid score stabilization and threshold clearance at 13-20 Mbp of accumulated data **(Figure 2B)**.

**Figure 2.**
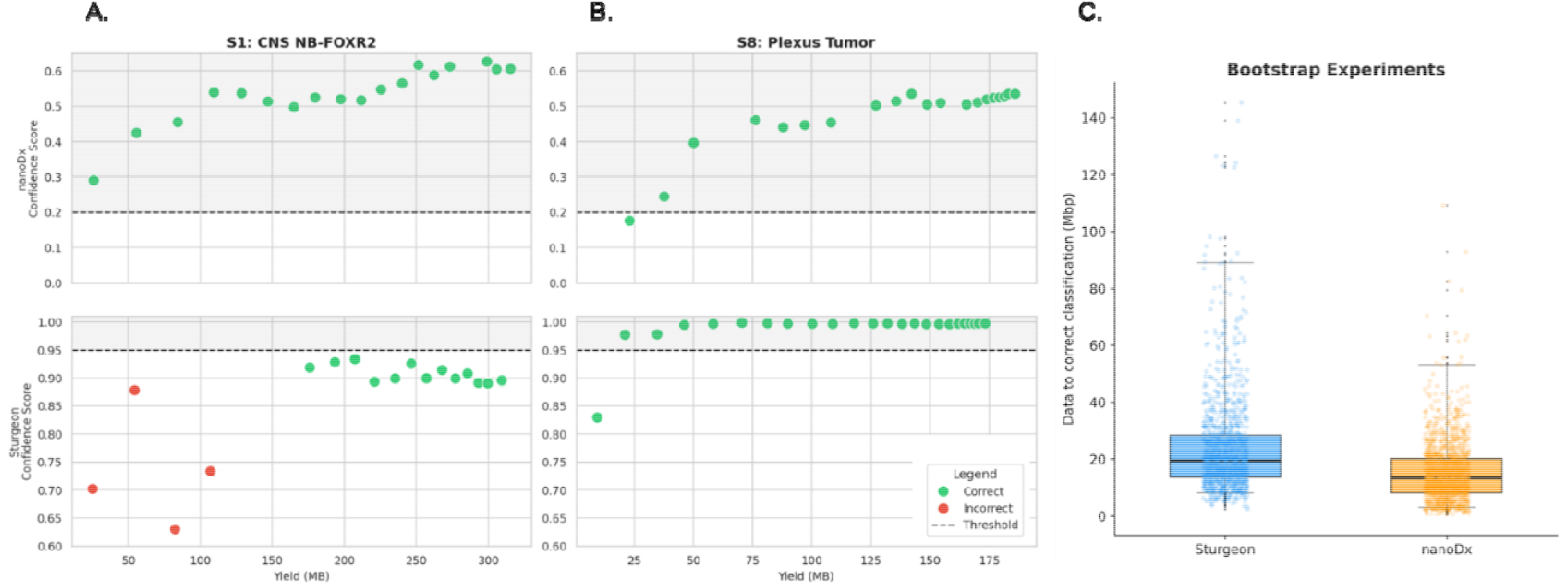
Longitudinal tracking of data buildup and bootstrap simulation of minimum data requirements. **(A, B)** Multi-panel tracking of intermediate data checkpoints over the course of active, continuous sequencing runs, showing how sequential data accumulation dynamically alters classifier assignments. (A) Data buildup tracking for Sample S1 (CNS NB-FOXR2, top: nanoDx; bottom: Sturgeon); under early data scarcity, insufficient data volume causes transient, incorrect assignments (red dots) by Sturgeon, which resolve into stable, correct, yet sub-threshold assignments as yield accumulates. Conversely, nanoDx crosses its analytical volume threshold early during sequencing. (B) Continuous data buildup tracking for Sample S8 (choroid plexus tumor, top: nanoDx; bottom: Sturgeon), demonstrating immediate score stabilization and robust alignment with ground truth as soon as initial, minimal data volumes are generated. **(C)** Bootstrap simulation of the minimum sequencing data required to reach correct classification, pooled across three samples (500 iterations per sample) for Sturgeon and nanoDx. Boxplots show the median (black line) and interquartile range of the data volume (Mbp) required for correct classification; whiskers extend to the 5th and 99th percentiles, and individual bootstrap replicates are overlaid as jittered points. Sturgeon required more data on average to converge on the correct classification than nanoDx.

To rigorously extend these observations and estimate data amounts required for significance classification, we performed an *in-silico* bootstrapping analysis **(Figure 2 C)**. Per-read CpG methylation calls were repeatedly sampled without replacement from each sequencing run, simulating 500 independent sequencing runs per sample. Among the samples selected for this analysis, we deliberately included the ATRT-MYC case (Sample S2), a methylation subtype known to pose a particular diagnostic challenge under sparse data conditions. Inter-subtype confusion between ATRT subgroups (MYC and SHH) has been independently documented in nanopore-based methylation classification under low-coverage settings^13^, making Sample S2 an informative test for classifier robustness at low data volumes. For each of the 500 bootstrap iterations per sample, we recorded the cumulative data yield at which each classifier first reached a correct, statistically significant assignment, converting the qualitative kinetic trends into a robust, quantitative distribution of throughput requirements for this sample.

For the tested ATRT-MYC sample, Sturgeon required a median of 19.4 Mbp of cumulative data to reach a confident, correct classification (score 19.4), and critically, produced zero false-positive classifications across all bootstrap iterations. NanoDx reached a correct classification at a lower median throughput of 19.4 Mbp (score 19.4). However, a subset of nanoDx bootstrap iterations exhibited a transient misclassification at low data volumes, defined as an incorrect class crossing the significance threshold before the correct class did. This behavior was most prominent in the ATRT-MYC sample, where early confusion with the molecularly related ATRT-SHH (ref acta) subgroup accounted for the majority of these events. Crucially, in every single instance, this early instability self-corrected as additional data accumulated, with the correct classification consistently emerging at a later checkpoint within the same bootstrap iteration. This pattern reinforces the core operational trade-off: nanoDx’s earlier sensitivity comes with a quantifiable, but transient and self-correcting, risk of premature misassignment, whereas Sturgeon’s higher threshold trades earlier readiness to eliminate this early risk entirely.

The present study demonstrates that low-cost ONT Flongle flow cells can support accurate methylation-based CNS tumor classification, even under conditions of extreme data sparsity, provided that data acquisition is guided by empirically defined thresholds. Despite substantial variability in sequencing yield, both Sturgeon and nanoDx achieved perfect concordance with neuropathological diagnoses when operating above significance thresholds, underscoring the robustness of current classification frameworks. However, their distinct performance characteristics highlight an important operational trade-off between early sensitivity and classification stability. Based on our bootstrap-derived estimates, we recommend implementing a minimum cumulative data threshold corresponding to the 99th percentile of data required for correct Sturgeon classification per Flongle run as a clinically actionable stopping criterion. At this level, cross-classifier concordance is maximized, transient nanoDx misclassifications are fully resolved, and Sturgeon achieves high-confidence assignments without false positives. Under these requirements, and given that additional tumor DNA is available, another flow cell may be used in order to complement missing data in cases where the initial sequencing run failed to yield confident classification. Adoption of this data-driven threshold and the Flongle flow cell may provide cost-effective, decentralized deployment of nanopore-based methylation diagnostics while maintaining the stringent reliability required for clinical decision-making.

## Data Availability

All data produced in the present study are available upon reasonable request to the authors

