## Supplementary material for "Towards Democratizing CNS Tumor Classification: 10-fold reduction in methylation sequencing cost with the Flongle Flow Cell": SI

Assaf Grunwald^1,2^, Galina Feinberg-Gorenshtein^3^, Helen Toledano^4,5^, Yehudit Birger^2,6^ ,Shai Izraeli^2,6^ and [Yuval Ebenstein](https://pubmed.ncbi.nlm.nih.gov/?term=Ebenstein+Y&cauthor_id=30485249)^1,5,7*^.

**Affiliations**

1. Department of Physical Chemistry, School of Chemistry, Tel Aviv University, Tel Aviv, Israel
2. Felsenstein Medical Research Center, Tel Aviv University, Petach Tikva, Israel
3. The Rina Zaizov Division of Pediatric Hematology-Oncology, Schneider Children’s Medical Center of Israel, Petach Tikva, Israel
4. Neuro-Oncology Unit, Dept. of Pediatric Hematology-Oncology, Schneider Children's Medical Center, Petah Tikva, Israel
5. The Gray Faculty of Medical and Health Sciences, Tel Aviv University, Tel Aviv, Israel
6. Faculty of Medicine, Tel Aviv University, Tel Aviv, Israel
7. Department of Biomedical Engineering, Faculty of Engineering, Tel Aviv University, Tel Aviv, Israel

**Clinical Cohort Ethics**

The study was conducted in accordance with the Declaration of Helsinki, under approval of The Rabin Medical Center (RMC) Institutional Review Board (IRB# 0012-08-RMC), approval 0039-17 RMC. Informed consent to participate in this study was obtained from patient parent or legal guardian. This study was approved by the relevant ethics committees, and all procedures were conducted in accordance with the 1964 Helsinki Declaration and its later amendments or comparable ethical standards.

**DNA Extraction and Quantification**

DNA from fresh-frozen tumor tissues was extracted using the QIAamp DNA Micro Kit (Qiagen, Hilden, Germany) according to the manufacturer’s instructions. For the FFPE samples, DNA was isolated as described before^1^ using either the QIAamp DNA FFPE Tissue Kit (Qiagen, Hilden, Germany) or the RecoverAll Multi-Sample RNA/DNA Kit (DNA component only; Invitrogen, Thermo Fisher Scientific, US), with the following modifications:

Instead of using xylene, tissue sections from 7–17 slides were pooled into a 1.5 mL tube containing 400 µL of digestion buffer. The tube was heated at 90 °C for 3 minutes, then centrifuged at 14,000 × g for 1 minute. A brief incubation on ice allowed the paraffin to solidify as a ring, which was then manually removed. This optimized heat-based deparaffinization protocol, routinely used in our clinical molecular oncology lab, reduces toxicity, simplifies handling, and ensures effective paraffin removal without compromising DNA recovery.

DNA concentrations were measured using a Qubit Flex fluorometer with the dsDNA HS Assay Kit (Invitrogen, Thermo Fisher Scientific, US).

**Modified Nanopore Library Preparation and Flongle Sequencing**

Library preparation for tumor-derived DNA was performed using the Rapid Sequencing Kit V14 (SQK-RAD114; Oxford Nanopore Technologies, UK) with specific modifications to optimize performance on low-input DNA. Crucially, specific reactions in the protocol were downscaled by 50% to accommodate smaller amounts of input DNA as follows: the DNA fragmentation mix volume was decreased to 5.5 µL by mixing 5 µL of purified, undiluted DNA with 0.5 µL of fragmentation mix. The adapter reaction was downscaled to a total volume of 2.5 µL by mixing 0.75 µL of “Rapid Adapter” with 1.75 µL of “Adapter Buffer (ADB)”. Then, 1 µL of this diluted adapter mix was added to the fragmentation mix. Finally, 5 µL of the adapter-DNA mix was combined with 15 µL of Sequencing Buffer (SB) and 10 µL of Library Beads (LIB) to be loaded onto the primed flow cell.

**Data Processing and CNS Classification**

**Sturgeon Classification Workflow:** Raw POD5 files were basecalled using the Dorado basecaller (Oxford Nanopore Technologies, UK) with the high-accuracy model configuration4.3.0 and the modified bases configuration4.3.0_5mCG_5hmCG@v1. Basecalled reads were aligned to the T2T-CHM13 reference genome using minimap2 v2.24^2^. The resulting BAM files were merged, sorted, and indexed using samtools v1.16^3^. To simulate the bisulfite sequencing features characteristic of the Capper reference training^4^, modkit v0.6. was utilized to convert 5hmC signals into 5mC signals and to extract single-CpG methylation *beta* values via the pileup command. The aggregated *beta* values were subsequently analyzed using the Sturgeon classifier under default parameters for central nervous system (CNS) tumor classification^5^.

**NanoDx Classification Workflow:** For nanoDx classification, pipelines were executed using version “nanoDx” 1.0rc3^6^ with default parameters, leveraging the pre-trained Capper et al. neural network model for CNS classification. Within this integrated framework, basecalling and methylation calling were performed via Dorado using the dna_r9.4.1_450bps_modbases_5hmc_5mc_cg_fast.cfg model, with alignments mapped using minimap2 v2.24^2^ against the hg19 reference genome.

**M-PACT Classification Workflow:** For M-PACT classification, POD5 files were basecalled as described above and aligned to the hg38 human reference genome. Single-CpG methylation values were extracted from the alignments using modkit v0.6.1 via the pileup command, isolating CpG sites (--cpg), combining modifications (--combine-mods), restricting calls to 5mC (--modified-bases 5mC), and aggregating across strands (--combine-strands). Classification was subsequently performed using the MethylVerse suite for CNS classification under default parameters^7^. Within this pipeline, the --impute and --regress configuration flags were explicitly enabled during runtime to facilitate deep-learning-based feature interpolation and regression adjustments under sparse data constraints.

**Longitudinal tracking of data buildup**: To evaluate the minimum sequencing data required for accurate intraoperative classification, we simulated a real-time sequencing scenario by processing Oxford Nanopore pod5 files incrementally across hourly timepoints. For each of the 12 samples, pod5 files were sorted chronologically and classified cumulatively, such that at timepoint h, all files sequenced up to and including hour h were provided as input to each classifier. Accumulated sequencing output at each timepoint was quantified from the aligned BAM using samtools flagstat. A classifier-specific confidence threshold was applied to determine a positive call (nanoDx: score ≥ 0.2; Sturgeon: score ≥ 0.95), and the earliest timepoint at which the predicted class matched the ground truth diagnosis above threshold was recorded as the time to correct classification.

**Bootstrap Simulation of Minimum Data Requirements**: To estimate the minimum sequencing throughput required for statistically confident classification, we performed a bootstrap resampling analysis for each classifier independently. Per-read CpG methylation calls were extracted from each sample's full BAM file using modkit extract calls (v0.4.2). For each of 500 independent iterations per sample, read IDs were randomly shuffled and subsampled without replacement in increments of 500 reads. At each increment, methylation calls were aggregated in memory and fed directly to the respective classifier — Sturgeon via modkit pileup-format aggregation followed by sturgeon inputtobed and sturgeon predict, and nanoDx via direct invocation of the NN classifier.predict() function from the nanoDx workflow — bypassing the Snakemake pipeline entirely. The cumulative sequencing yield (in Mbp) at the step where the correct class first achieved a statistically significant score (Sturgeon ≥ 0.95; nanoDx ≥ 0.20) was recorded as the classification threshold for that iteration. Iterations in which the correct class did not reach significance within the full dataset were recorded as unclassified. All scripts are available upon request.

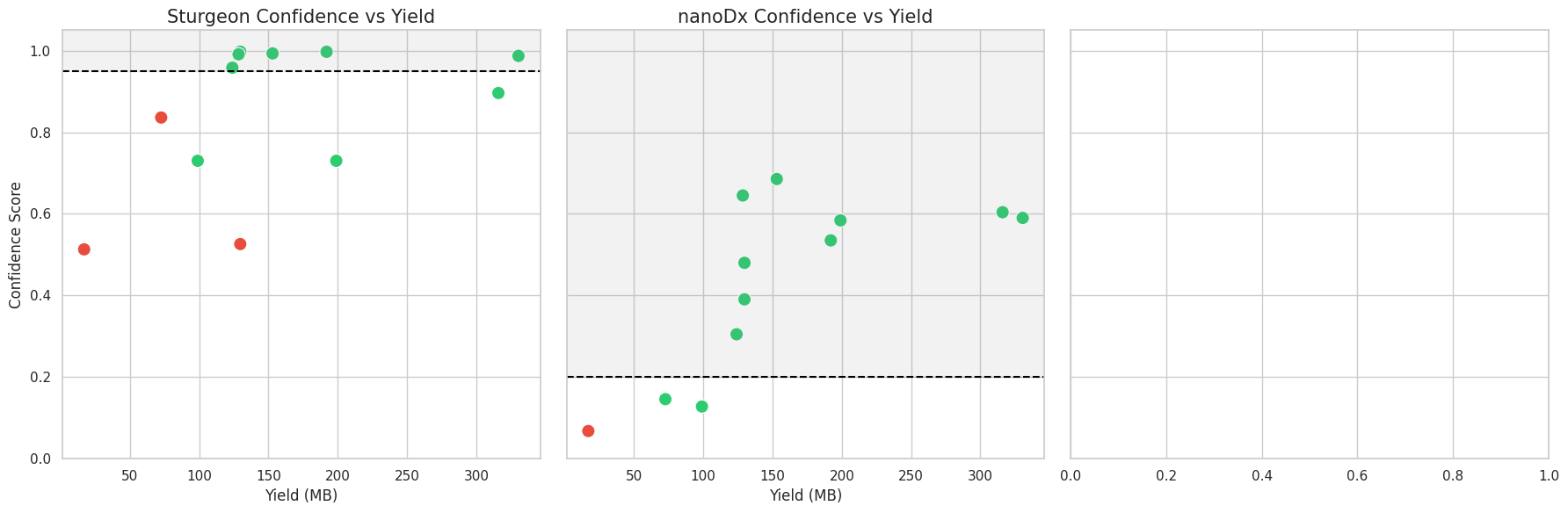

***Figure 2: Final classification confidence scores as a function of total basecalled data across the 12-run Flongle cohort.*** *Side-by-side scatter plots displaying the overall cohort endpoints for Sturgeon (left panel) and nanoDx (right panel) across cumulative data yields (Mbp). Each point represents an individual Flongle sequencing run, color-coded by diagnostic concordance with the neuropathologist-confirmed*
